# Association between fruit and vegetable consumption and chronic diseases among food pantry users

**DOI:** 10.1101/2024.03.14.24304291

**Authors:** Jiacheng Chen, Akiko S. Hosler, Thomas J. O’Grady, Xiaobo X. Romeiko

## Abstract

**Introduction:** Fruit and vegetable (FV) consumption can be a protective factor for chronic diseases, but few studies have investigated FV’s impact on health in the context of food/nutrition assistance system.

**Methods:** We used three health survey data collected in Upstate New York communities to construct a predictive model of food pantry use. The model was applied to a Northeastern US regional subset of SMART Behavioral Risk Factor Surveillance System (BRFSS) data to identify potential food pantry users. The associations between FV intake and diabetes, hypertension, and BMI were examined through multivariable logistic regression and linear regression analyses with food pantry use as a potential effect modifier.

**Results:** The analysis dataset had 5,257 respondents, and 634 individuals were estimated as food pantry users. Consumption of vegetables was associated with decreased odds of hypertension and a lower BMI regardless of food pantry use. Consumption of fruits was associated with decreased odds of diabetes regardless of food pantry use. The association between fruit consumption and BMI was modified by food pantry use. Among food pantry users, consumption of fruits was associated with a greater BMI, while among food pantry non-users, it was associated with a lower BMI.

**Conclusion:** The overall protective effects of increased FV consumption on chronic diseases suggest that increasing FV availability in food pantries may not only alleviate hunger but also improve health. Further research is needed to investigate the role of fruit including 100% fruit juice consumption and BMI among food pantry users.

## Introduction

Chronic diseases such as hypertension, diabetes, and obesity impose considerable burdens on people and the healthcare systems in the US^1^ and around the globe^2^. While these chronic diseases have multiple causal pathways and risk factors^3–6^, there are also protective factors. Consumption of fruits and vegetables (FVs) is one of the protective factors that are associated with improved blood pressure, and lower A1C and BMI^7–11^. The levels of FV consumption, however, vary across US population^12^. Individuals who are food insecure consume fewer amounts of FVs and have higher prevalence of chronic diseases compared to people who are food secure^13^.

Food pantries were initially created to provide temporary nutritional relief to individuals experiencing food insecurity, but they have become integral to ensuring access to high-quality nutritious food among those in need^14,15^. Interventions to increase the availability of FVs in food pantries have shown improvements in some dietary and health behaviors among their clients^16,17^. However, more evidence is needed to show that FV consumption can improve chronic disease outcomes in the context of the food/nutrition assistance system. A scarcity of large population-based health datasets with food pantry usage and health outcomes information has been a primary obstacle to conducting research in this area.

This study seeks to address these research gaps by utilizing multiple existing datasets. First, we developed a predictive model of food pantry use using demographic and spatial variables from existing community survey datasets. Next, we applied the predictive model to a larger, regionally representative health dataset, and examined the associations between FV consumption and hypertension, diabetes, and BMI with food pantry use as a potential effect modifier. Information generated from this study will inform regional nutrition and public health professionals about potential health benefits of increased FV availability in food pantries.

## Methods

### Study Community

The study community is five Metropolitan Statistical Areas (MSAs) in the Northeastern US. Specifically, the Albany-Schenectady-Troy MSA, the Buffalo-Cheektowaga MSA, the Rochester MSA in New York State (NYS), the Burlington-South Burlington MSA in Vermont, and the Springfield MSA in Massachusetts. These MSAs were selected based on the urbanicity and location (small- to mid-sized cities in Northeaster US), population size (less than 1.2 million), climate and vegetation (USDA plant hardiness zone 4 or 5), and the availability of SMART Behavioral Risk Factor Surveillance System (BRFSS) data. SMART BRFSS is a project by the Centers for Disease Control and Prevention to compile BRFSS telephone health survey data for MSAs and other types of metropolitan divisions with at least 500 completed interviews. Figure 1 shows the locations of the five selected MSAs.

**Figure 1.**
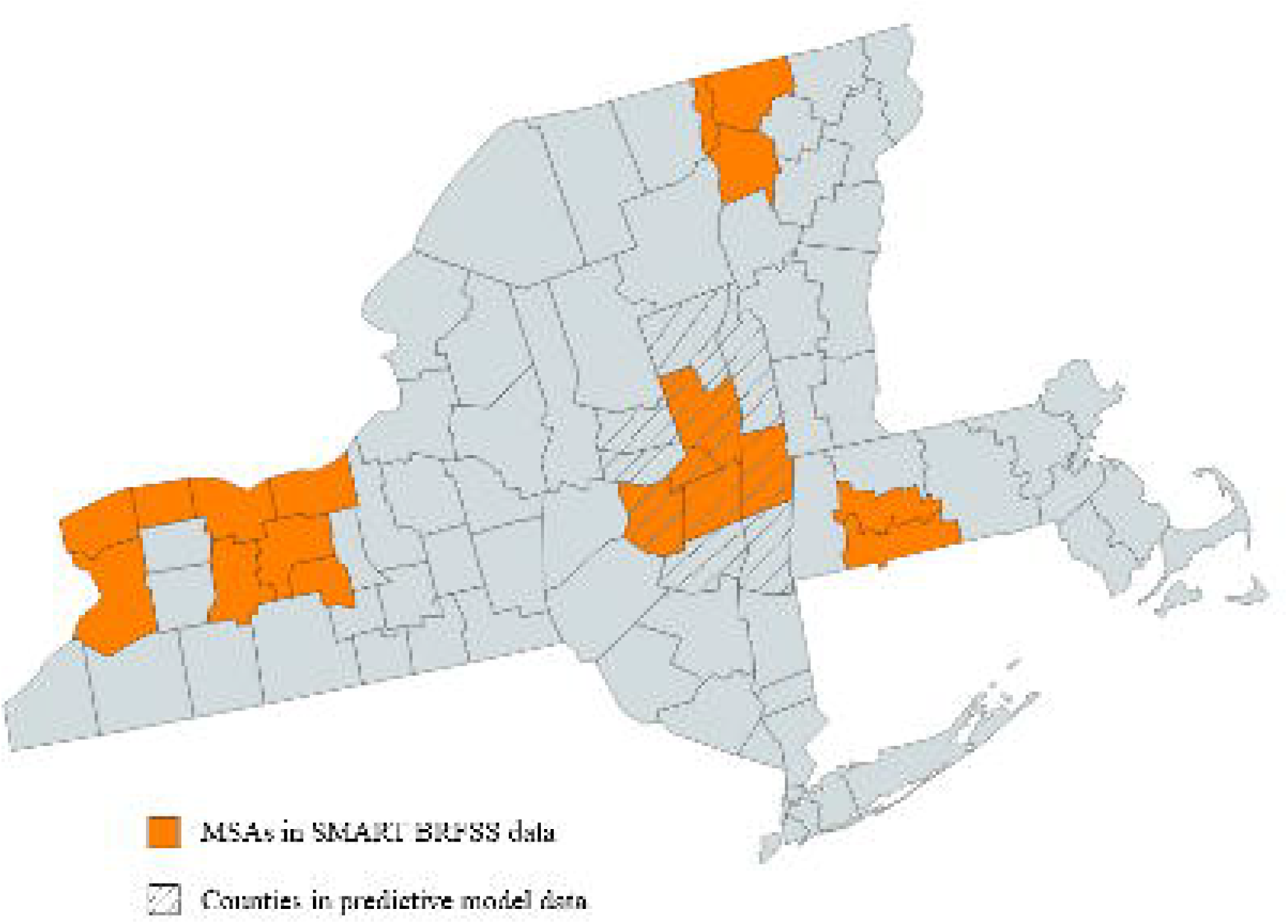
Five Metropolitan Statistical Areas (MSAs) included in the SMART BRFSS dataset and New York State counties where data for the predictive model were collected

### Food Pantry User Predictive Model Analysis

The BRFSS surveys, the source of this study’s main data, do not collect food pantry use information. Therefore, our first step was to build a predictive model for food pantry use using three existing survey data from Upstate NYS communities. UMatter Schenectady (UMS) was a health interview survey conducted in Schenectady County in 2014^18^. Food Access Survey 1 (FAS-1) and Minority Health Disparity Survey (MHDS) were online surveys of residents in 11 counties in 2020-2021^19^. MHDS was introduced to augment representations of low-income racial/ethnic minorities and rural residents in our food access research, and it was administered partially concurrently with FAS1. These three datasets contained comparable demographic, spatial, and food pantry usage information collected from English- or Spanish-speaking adults 18 years and older. The sample size of UMS, FAS1, and MHDS were 2,234, 595 and 454, respectively. Table 1 describes characteristics of survey data in this study, and figure 1 shows the locations of the survey communities.

**Table 1.** Characteristics of the survey datasets used in this study.

| Dataset | Year | Sample Size | Geographic Coverage | Variable Availability |  |  |  |
| --- | --- | --- | --- | --- | --- | --- | --- |
|  |  |  |  | Fruit & vegetable | Chronic disease | Food pantry | Demo-graphics |
| UMS | 2014 | 2,234 | 1 county in NY | Limited (1 question) | **Yes** | **Yes** | **Yes** |
| FAS1 | 2020-21 | 595 | 11 counties in NY | **Yes** (2 questions) | No | **Yes** | **Yes** |
| MHDS | 2021 | 454 | 11 counties in NY | **Yes** (2 questions) | No | **Yes** | **Yes** |
| SMART BRFSS | 2015 | 5,454 | 3 MSAs in NY, 1 MSA in VT, 1 MSA in MA | **Yes** (6 questions) | **Yes** | No | **Yes** |

We combined the three datasets and constructed a multivariable logistic regression model for food pantry use. Predictor variables were several demographic and spatial variables commonly shared across these datasets and SMART BRFSS. We converted the county variable into a 6-level urbanicity variable derived from the NCHS Urban-Rural Classification Scheme for Counties^20^.

We applied a backward-deletion strategy and used AIC, likelihood ratio tests, and the ROC curve to examine the model fit. The final model contained education, race/ethnicity, household income, and urbanization scale as predictors. The output of the predictive model had a value from 0 to 1. To find the optimal cut-point to identify food pantry users, we employed Youden’s J statistic based on the ROC curve. The optimal cut-point is the threshold that maximizes the distance to the identity line, which is also the point at which the sum of sensitivity and specificity is maximized.

### SMART BRFSS Associative Model Analysis

The goal of the associative model analysis was to assess the associations between FV consumption and three chronic disease indicators and examine whether food pantry use would modify the associations. We obtained the 2015 SMART BRFSS data subsets for the Albany-Schenectady-Troy, Buffalo-Cheektowaga, Rochester, Burlington-South Burlington, and Springfield MSAs. In all BRFSS surveys, fruit and vegetable questions were administered in odd years. 2015 data were selected because it was the last and most recent data collection year when vegetable consumption was assessed by nutrient groups.

Fruit consumption was measured by two questions asking about times 100% fruit juice and fruits were taken in the last 30 days. Vegetable consumption was measured by four questions asking about times beans, dark green vegetables, orange-colored vegetables, and other vegetables including 100% vegetable juice were taken in the last 30 days. All forms of fruits and vegetables including cooked or raw, fresh, frozen, or canned were considered. After obtaining frequency measures of FV consumption, we used the linear regression model developed by Moore et al. to estimate the daily consumption amount in cups^21^. This model included age, sex, race/ethnicity, and poverty-income ratio as factors to estimate FV consumption in cups. In the analysis, age and sex served as effect modifiers, while other variables acted as predictors.

Chronic disease outcomes were hypertension and diabetes both based on self-reported diagnosis (yes/no), and BMI (kg/m^2^) which was computed using self-reported height and weight. We estimated food pantry use by applying the predictive model described in the previous section. We constructed logistic regression models for diabetes and hypertension, and linear regression models for BMI, with fruit and vegetable consumption separately. We included age groups, sex, employment status, physical activity, smoking, and alcohol consumption as covariates in the initial models. To avoid overadjustment, we excluded the variables used as predictors in the food pantry use predictive model^22^.

We employed a backward-deletion strategy to incrementally exclude covariates with large p-values. We used the interaction terms “food pantry use*vegetable consumption” and “food pantry use*fruit intake” to examine potential effect modification of food pantry use. If a significant (p<0.01) effect modification was identified, we stratified the health impact of FV consumption for food pantry users and non-users.

This study was conducted according to the guidelines laid down in the Declaration of Helsinki, and all procedures involving research study participants were approved by the University at Albany Institutional Review Board.

## Results

In the predictive model analysis, 3,227 complete cases were included in the combined dataset. Low education level, racial/ethnic minority status, living in areas with a higher urbanization level, having lower household incomes or not responding to the income question were positively associated with food pantry use. The optimal cut-point from the ROC curve was 0.177.

The analysis dataset of SMART BRFSS had 5,257 respondents. When we applied the predictive model to the dataset, 634 individuals (12.1%) were identified as potential food pantry users. The demographic characteristics of SMART BRFSS respondents are described in table 2. Briefly, 68.3% were working-age adults (age <65 years), 57.6% were women, and 85.8% were non-Hispanic Whites.

**Table 2.** Characteristics of the analysis dataset of SMART BRFSS (N=5,257)

| Characteristic |  | N | % |
| --- | --- | --- | --- |
| Age (years) | 18-34 | 811 | 15.4 |
|  | 35-54 | 1,527 | 29.0 |
|  | 55-64 | 1,252 | 23.8 |
|  | 65 and older | 1,667 | 31.7 |
| Sex | Male | 2,227 | 42.4 |
|  | Female | 3,030 | 57.6 |
| Race/Ethnicity | Hispanic | 242 | 4.6 |
|  | Black, not Hispanic | 268 | 5.1 |
|  | White, not Hispanic | 4,511 | 85.8 |
|  | Asian | 91 | 1.7 |
|  | Multiple Races/Other | 145 | 2.8 |
| Education | High school or less | 1,718 | 32.7 |
|  | Some college or Associate degree | 1,319 | 25.1 |
|  | College degree or advanced | 2,220 | 42.2 |
| Household Income | Less than \$10,000 | 195 | 3.7 |
| | \$10,000 - 49,999 | 1,851 | 35.2 |
| | \$50,000 or more | 2,361 | 44.9 |
|  | Unreported | 850 | 16.2 |
| Employment | Employed | 2,771 | 52.7 |
|  | Unemployed | 243 | 4.6 |
|  | Not in the labor force | 2,242 | 42.6 |
| MSA | Albany-Schenectady-Troy, NY | 888 | 16.9 |
|  | Rochester, NY | 756 | 14.4 |
|  | Buffalo-Niagara Falls, NY | 722 | 13.7 |
|  | Burlington, VT | 1,746 | 33.2 |
|  | Springfield, MA | 1,145 | 21.8 |
| Food pantry use (estimated) | Yes | 634 | 12.1 |
|  | No | 4,623 | 87.9 |

In the associative model analysis, we found that food pantry use was not an effect modifier for the association between FV consumption and hypertension or diabetes. Fruit consumption had no significant effect on hypertension; however, one cup of daily vegetable consumption decreased the odds of hypertension by 21% (OR: 0.79, 95% CI: 0.70-0.89) in the final model, regardless of food pantry use (table 3). For diabetes outcome, one cup of daily fruit intake was also protective, with a 17% reduction of the odds of diabetes (OR: 0.83, 95% CI: 0.69-0.99) in the final model regardless of food pantry use. Vegetable intake had no significant effect on diabetes.

**Table 3.** Multivariable logistic regression models for hypertension and diabetes.

|  |  | Hypertension<br>OR (95% CI) | Diabetes<br>OR (95% CI) |
| --- | --- | --- | --- |
| Daily Fruit Consumption<br>(Cup) | Crude Model | 0.96 (0.86, 1.06) | 0.83 (0.70, 0.99) |
|  | Final Model<sup>a</sup> | 0.96 (0.86, 1.08) | **0.83** (0.69, 0.99) |
| Daily Vegetable<br>Consumption (Cup) | Crude Model | 0.53 (0.47, 0.59) | 0.59 (0.49, 0.70) |
|  | Final Model<sup>a</sup> | **0.79** (0.70, 0.89) | 0.92 (0.76, 1.09) |

a. Final model is adjusted for age groups, smoking status, alcohol consumption and physical activity.

**Table 4.**
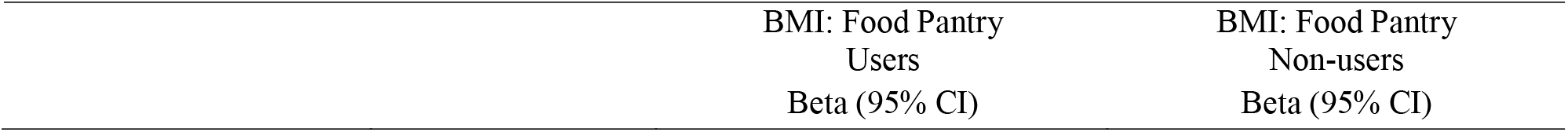
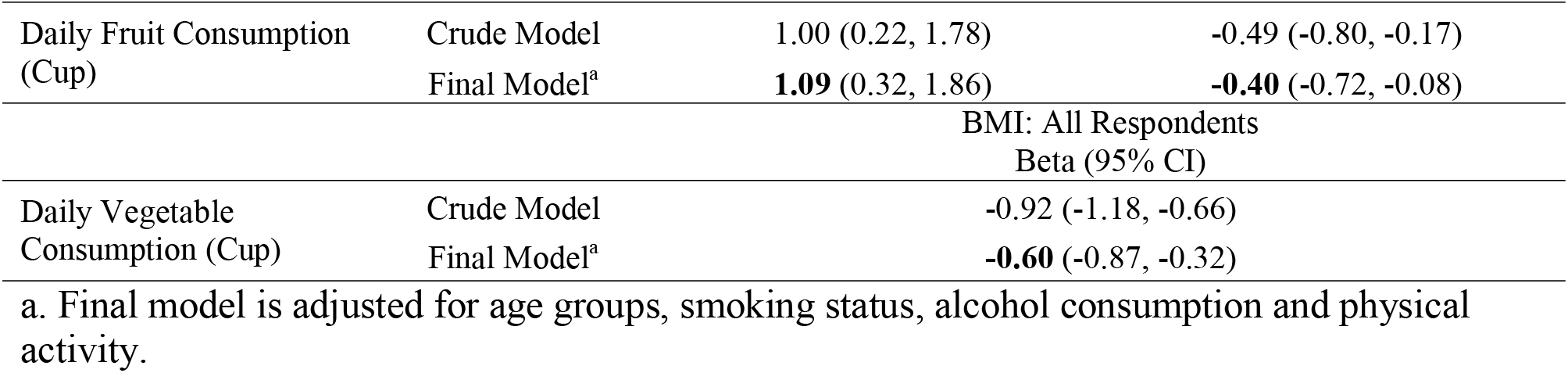
Multivariable linear regression models for BMI (Kg/m^2^)

For the association between FV consumption and continuous BMI, we found that food pantry use was an effect modifier for fruit consumption only. In the covariate-adjusted final models, one cup of daily fruit consumption increased BMI by 1.09 kg/m^2^ (95% CI: 0.32, 1.86) among food pantry users, while it decreased BMI by 0.40 kg/m^2^ (95% CI: -0.72, -0.08) among food pantry non-users. Vegetable consumption had a universally protective effect on BMI. One cup of daily vegetable consumption decreased BMI by 0.6 kg/m^2^ (95% CI: -0.8, 7-0.32) in the final model regardless of food pantry use.

## Discussion

This study investigated potential health impacts of FV consumption on hypertension, diabetes, and BMI using a regionally representative large health survey dataset. Food pantry use was estimated by applying a predictive model generated from three community survey datasets from the same region. This analytic technique allowed us to examine whether food pantry use would modify the FV consumption and chronic disease relationship.

Overall, this study provided evidence that FV consumption has protective associations with chronic disease outcomes. Increasing daily consumption of fruit or vegetables by a cup could decrease the odds of diabetes or hypertension and decrease BMI. This study also demonstrated that food pantry use did not modify the FV consumption and chronic disease relationship, except for the association between fruit consumption and BMI. The positive association between fruit consumption and BMI among food pantry users has not been reported elsewhere. Additional investigation revealed that food pantry users had higher BMI (Mean 30.07 kg/m^2^, SD 8.19 kg/m^2^) compared to food pantry non-users (Mean 27.48 kg/m^2^, SD 5.86 kg/m^2^), and the t-test comparing the means of the two groups yielded a significant difference (p<0.001). Additionally, food pantry users had a lower amount of total daily fruit consumption (1.30 cups vs 1.51 cups), but a higher consumption of 100% fruit juice (0.51 cup vs 0.37 cup) compared to food panty non-users. Although 100% fruit juice may contain as much sugar as regular soft drink, there is no conclusive evidence that 100% fruit juice increases BMI in US adults^23^. It is also argued that 100% fruit juice provides important vitamins, minerals, and dietary bioactives that contribute to overall health^24^. Further investigation is warranted for the association between fruit consumption and BMI in food pantry users.

This study is not without limitations. Methodologically, the use of regression model-created variables can introduce Berkson bias^25^, a random error that reduces statistical power. However, the sample size for the associative model analysis (5,227) was large enough to overcome the statistical power reduction that may have been introduced by Berkson bias. Food pantry use was estimated and not actually measured. The prediction model included only demographic and urbanicity variables. Adding variables for Food and Nutrition Assistance Program (SNAP) participation and use of group meal sites would significantly improve the overall prediction model fit^26^. However, this was not feasible because SMART BRFSS does not collect such information. Because food pantry usage does not have eligibility criteria or membership registration requirements, “food pantry users” is a fluid concept. Nonetheless, the proportion of food pantry users in this study (12.1%) was very close to the estimated percentage of America households experiencing food insecurity (12.5%) in 2015^27^. Finally, the cross-sectional design prevents establishing causality between FV intake and chronic disease risks.

Although fruit consumption is associated with a small increase in BMI among food pantry users, the overall health benefit of increasing FV consumption is much greater. FVs provided by food pantries represent an important way to alleviate hunger and food insecurity while also contributing to better health among food pantry users. This study provides important evidence to food pantries on the benefits of increasing the availability of FVs in food pantries and encouraging the consumption of FVs among food pantry users.

## Data Availability

The UMS, FAS1, MHDS data in the present study are available upon reasonable request to the authors. The SMART BRFSS2015 data is available online at: https://www.cdc.gov/brfss/smart/smart_2015.html

## Acknowledgements

This work was supported by the Foundation for Food and Agriculture Research (FFAR; 557409). FFAR had no role in the design, analysis, or writing of this article. We thank Drs. Beth Feingold, Roni Neff, and Christine Bozlak for their support of data collection efforts. (2027 words)

